# The effect of international travel restrictions on internal spread of COVID-19

**DOI:** 10.1101/2020.07.12.20152298

**Authors:** Timothy W Russell, Joseph T Wu, Sam Clifford, CMMID COVID-19 working group, W John Edmunds, Adam J. Kucharski, Mark Jit

## Abstract

**Background:** Countries have restricted international arrivals to delay the spread of COVID-19. These measures carry a high economic and social cost. They may have little impact on COVID-19 epidemics if there are many more cases resulting from local transmission compared to imported cases.

**Methods:** To inform decisions about international travel restrictions, we compared the ratio of expected COVID-19 cases from international travel (assuming no travel restrictions) to the expected COVID-19 cases arising from internal spread on an average day in May 2020 in each country. COVID-19 prevalence and incidence were estimated using a modelling framework that adjusts reported cases for under-ascertainment and asymptomatic infections.

**Findings:** With May 2019 travel volumes, imported cases account for <10% of total incidence in 103 (95% credible interval: 76 − 130) out of 142 countries, and <1% in 48 (95% CrI: 9 − 95). If we assume that travel would decrease compared to May 2019 even in the absence of formal restrictions, then imported cases account for <10% of total incidence in 109-123 countries and <1% in 61-88 countries (depending on the assumptions about travel reductions).

**Interpretation:** While countries can expect infected travellers to arrive in the absence of travel restrictions, in most countries these imported cases likely contribute little to local COVID-19 epidemics. Stringent travel restrictions may have limited impact on epidemic dynamics except in countries with low COVID-19 incidence and large numbers of arrivals from other countries.

**Funding:** Wellcome Trust, UK Department for International Development, European Commission, National Institute for Health Research, Medical Research Council, Bill & Melinda Gates Foundation

**Research in context:** *Evidence before this study:* Countries are at different stages of COVID-19 epidemics, so many have implemented policies to minimise the risk of importing cases via international travel. Such policies include border closures, flight suspensions, quarantine and self-isolation on international arrivals. Searching PubMed and MedRxiv using the search: (“covid” OR “coronavirus” OR “SARS-CoV-2”) AND (“travel” OR “restrictions” OR “flight” OR “flights” OR “border”) from 1 January – 10 July 2020 returned 118 and 84 studies respectively, of which 39 were relevant to our study. These studies either concentrated in detail on the risk of importation to specific countries or used a single epidemiological or travel dataset to estimate risk. Most of them focused on the risk of COVID-19 introduction from China or other countries with cases earlier in 2020. No study combined country-specific travel data, prevalence estimates and incidence estimates to assess the global risk of importation relative to current local transmission within countries.

*Added value of this study:* We combined data on airline passengers and flight frequencies with estimates of COVID-19 prevalence and incidence (adjusted for underreporting and asymptomatic cases), to estimate the risk of imported cases, relative to the level of local transmission in each country. This allows decision makers to determine where travel restriction policies make large contributions to slowing local transmission, and where they have very little overall effect.

*Implications of all the available evidence:* In most countries, imported cases would make a relatively small contribution to local transmission, so travel restrictions would have very little effect on epidemics. Countries where travel restrictions would have a large effect on local transmission are those with strong travel links to countries with high COVID-19 prevalence and/or countries which have successfully managed to control their local outbreaks.

## Introduction

Coronavirus disease 2019 (COVID-19) is an illness caused by Severe Acute Respiratory Syndrome coronavirus 2 (SARS-CoV-2), which was first detected in Wuhan, China, in late 2019. Since then, it has been spread by travellers to almost every country in the world, and was declared a pandemic by the World Health Organization on 11 March 2020 ^1^. In the absence of effective pharmaceutical measures for prevention and treatment, countries have imposed a range of response measures to delay the spread of SARS-Cov-2, and hence enable health systems to cope with the expected sharp rise in health care demand.

One such intervention that has been widely used is international travel restrictions. Early travel restrictions focused on countries with early outbreaks (like China, Iran and Italy) but as SARS-CoV-2 spread to more countries, the list of origin countries on countries’ travel restriction lists has grown. The World Tourism Organization reports that every country in the world had imposed some form of COVID-19-related travel restriction by 20 April 2020, the most extensive travel restrictions in history ^2^. However, which restrictions were implemented differs from country to country and include border closures, flight suspensions, quarantine and self-isolation. Measures may also be applied indiscriminately or targeted at specific places of origin.

International travel restrictions carry a high economic and social cost. Much of global tourism, trade, business, education and labour mobility relies on cross-border movement of people. The United Nations Conference on Trade and Development (UNCTAD) estimates that the world’s tourism sector will lose value worth 1.6-2.8% of global gross domestic product as a result of COVID-19; this excludes the value of lost non-tourism travel ^3^. The social cost comes in the form of lost opportunities for family and friend reunion, international education and career development. According to International Health Regulations (2005) travel restrictions “shall not be more restrictive of international traffic and not more invasive or intrusive to persons than reasonably available alternatives that would achieve the appropriate level of health protection.” ^4^. Hence there are strong economic, humanitarian and legal reasons to only impose international travel restrictions when the benefits outweigh the costs.

Travel restrictions have clear benefits when there are zero or few cases in the destination country. For instance, restrictions on travellers from Wuhan, or China more generally, in early 2020 may have contributed to slowing global spread of SARS-CoV-2 ^56^. However, once case numbers within a country are sufficiently large that local outbreaks have been established and are self-sustaining, travel restrictions become less effective. For instance, the ban on European travellers to the United States on 12 March 2020 was too late to prevent a large epidemic in New York that already had been seeded mainly by European travellers ^7^. Countries with established epidemics attempting to reduce COVID-19 incidence through stringent physical distancing measures such as lockdowns may impose travel restrictions to accelerate the reduction of new cases. However, this would only be effective if the number of cases being imported from international travellers would contribute substantially to overall incidence. Hence decisions around travel restrictions are complex; they need to take into account local transmission, COVID-19 prevalence in source countries of travellers and the volume of travel from those countries.

In this paper, we provide information to countries about the potential benefit of international travel restrictions by comparing the number of cases resulting from international travel to those resulting from local transmission in 142 countries.

## Methods

### Prevalence by country

Our analysis combines estimates of SARS-CoV-2 prevalence and incidence for each country with detailed flight data. The prevalence and incidence estimates are derived using statistical modelling methods described elsewhere ^8^ and summarised here. First, the level of case ascertainment in each country is estimated using the ratio of a delay-adjusted country specific CFR and an assumed published “baseline” CFR ^9^. Then, temporal variation in under-ascertainment is inferred using a Gaussian Process framework. Finally, these temporal under-ascertainment estimates are used to adjust the confirmed case time series ^10^. The adjusted case data represent the estimated true number of symptomatic individuals in each country, which are typically significantly larger than the confirmed case numbers ^8,11^.

Incidence is estimated as the number of new cases on the most recent day that the country in question had new cases, after adjusting for under-ascertainment and asymptomatic infections. Prevalence on each day is estimated as the sum of the new cases over the nine most recent days, i.e. assuming an infectious period of ten days ^12–14^. This is then converted to a proportion by dividing by the country’s population.

### International travellers

International travel has reduced greatly since the COVID-19 pandemic began ^2^ because of travel restrictions, but also because individual self-exclusion due to fear of infection and reduced business and tourism opportunities. Hence we considered four scenarios for international travel in May 2020 in the hypothetical case that there were no travel restrictions:

Scenario A: The number of travellers between each country was estimated using the number of passengers booked on flights, using data from the Official Aviation Guide (OAG) in May 2019, i.e. assuming that travel patterns in 2020 would be identical to 2019 in the absence of travel restrictions.

Scenario B: The OpenSky dataset provides data on the number of flights each day between pairs of countries. We adjusted the number of international travellers between countries downwards using the ratio of the number of flights in the OpenSky database in May 2019 and May 2020. This gave a mean reduction of 69% (range 0% − 99%) across countries (Figure 3). Where data were not available, we applied the mean reduction across pairs of countries with data.

Scenarios C and D: The number of flights do not completely capture the reduction in passengers, since aircraft occupancy has also decreased in 2020. The International Air Transport Association (IATA) projects that passenger departures will decline by 50.6% in 2020 compared to 2019 ^15^. In the absence of travel restrictions, we assumed two scenarios about international travellers: 25% reduction (scenario C) and 50% reduction (scenario D).

### Imported cases

The number of cases imported from a source country to a destination country on a particular date is estimated as the product of the prevalence on that date in the source country multiplied by the number of travellers from that country to the destination country on a single day in May 2020. The total number of imported cases on that date is then estimated by summing the cases imported from all countries with travellers to the destination country. We then calculated the ratio of imported cases to total incidence.

For countries where imported cases are predicted to account for over 1% of local incidence, we estimated the proportion of incoming travellers that needed to be averted in order to bring this proportion below 1%. We assumed that incoming travellers would be averted in order of COVID-19 prevalence in their countries of origin, i.e. averting travellers from the highest prevalence country first.

## Results

Figure 1 shows the risk rating for each country, based on the ratio of imported cases to total incidence, for different scenarios about passenger reductions in May 2020. Even in the worst-case scenario of no change in travel patterns compared to May 2019 in the absence of travel restrictions (Figure 1, Panel A), we estimate that in 109 countries out of 142 modelled imported cases contribute to <10% of local incidence.

**Figure 1.**
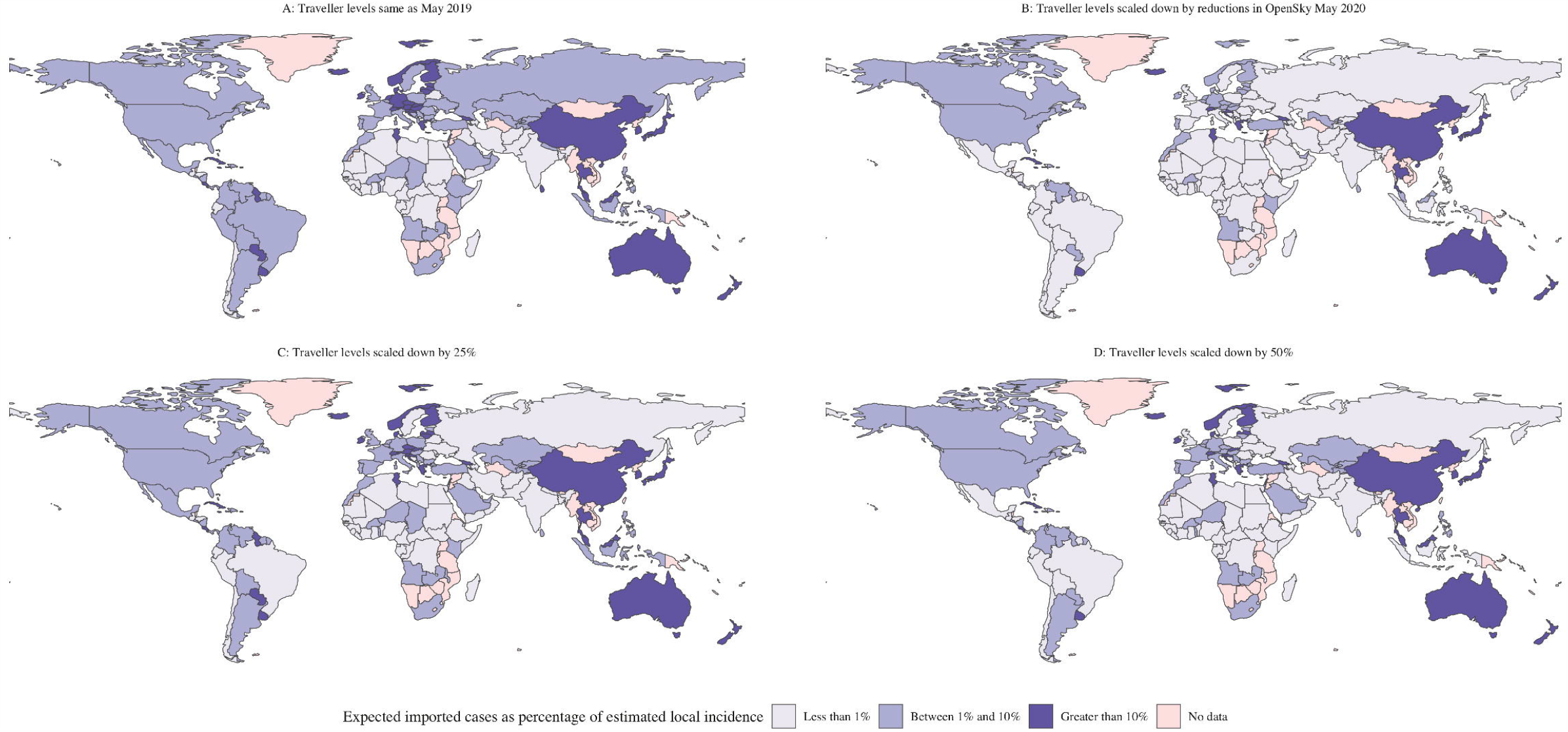
Risk rating by country, in the absence of international travel restrictions, in each of the four scenarios about international travellers in May 2020. (A) Travel assumed to be at the same levels as May 2019. (B) Traveller numbers scaled downwards based on the reduction in flights in May 2020 reported by OpenSky. (C) Traveller numbers scaled down by 25%. (D) Traveller numbers scaled down by 50%.

There is little change in country risk ratings when we assume that international travellers in 2020 decreased compared to 2019. In particular, most countries where imported cases are estimated to contribute to 10% or more of local incidence (including China, Thailand and Australia) remain in that category across all scenarios. The scenarios assuming reduction in travel have the largest effect on European and Latin American countries, where imported cases are estimated to fall below 1% of local incidence as assumed travellers to those regions decrease.

Figure 2 shows the proportion by which international arrivals need to be averted to bring imported cases to below 1% of local incidence, in all countries in scenario B where it is over 1%. Most of these countries would need to avert the majority of their international arrivals, although there are a few that may be able to bring imported cases to below 1% of local incidence by averting less than a quarter of arrivals. Figure 3 shows the relationship between imported cases and local outbreak size.

**Figure 2.**
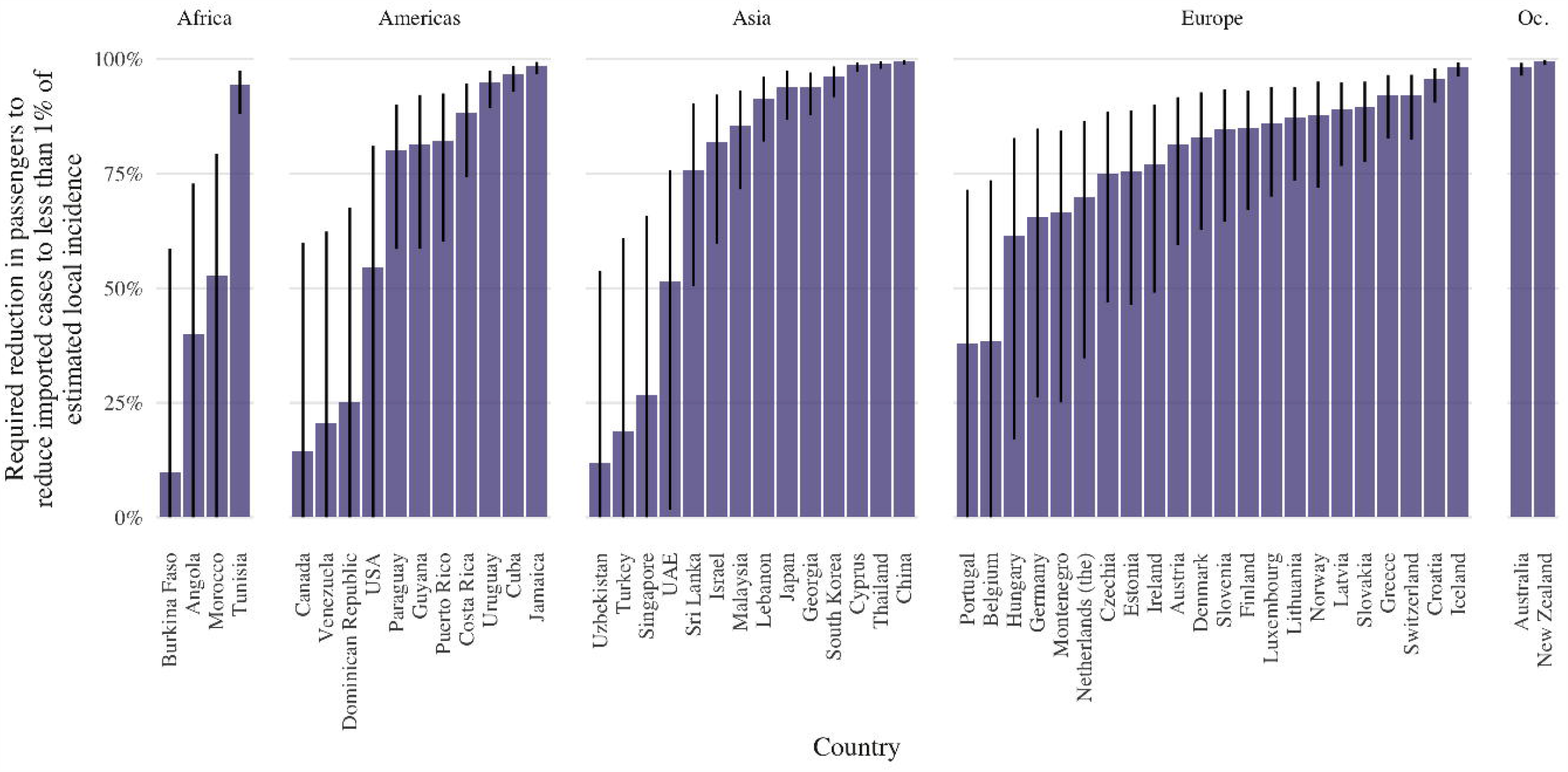
Percentage reduction in passenger numbers required for countries in scenario B where imported cases account for more than 1% of local incidence to bring that proportion below 1%. Countries are grouped by United Nations Region (Oc. is Oceania).

**Figure 3.**
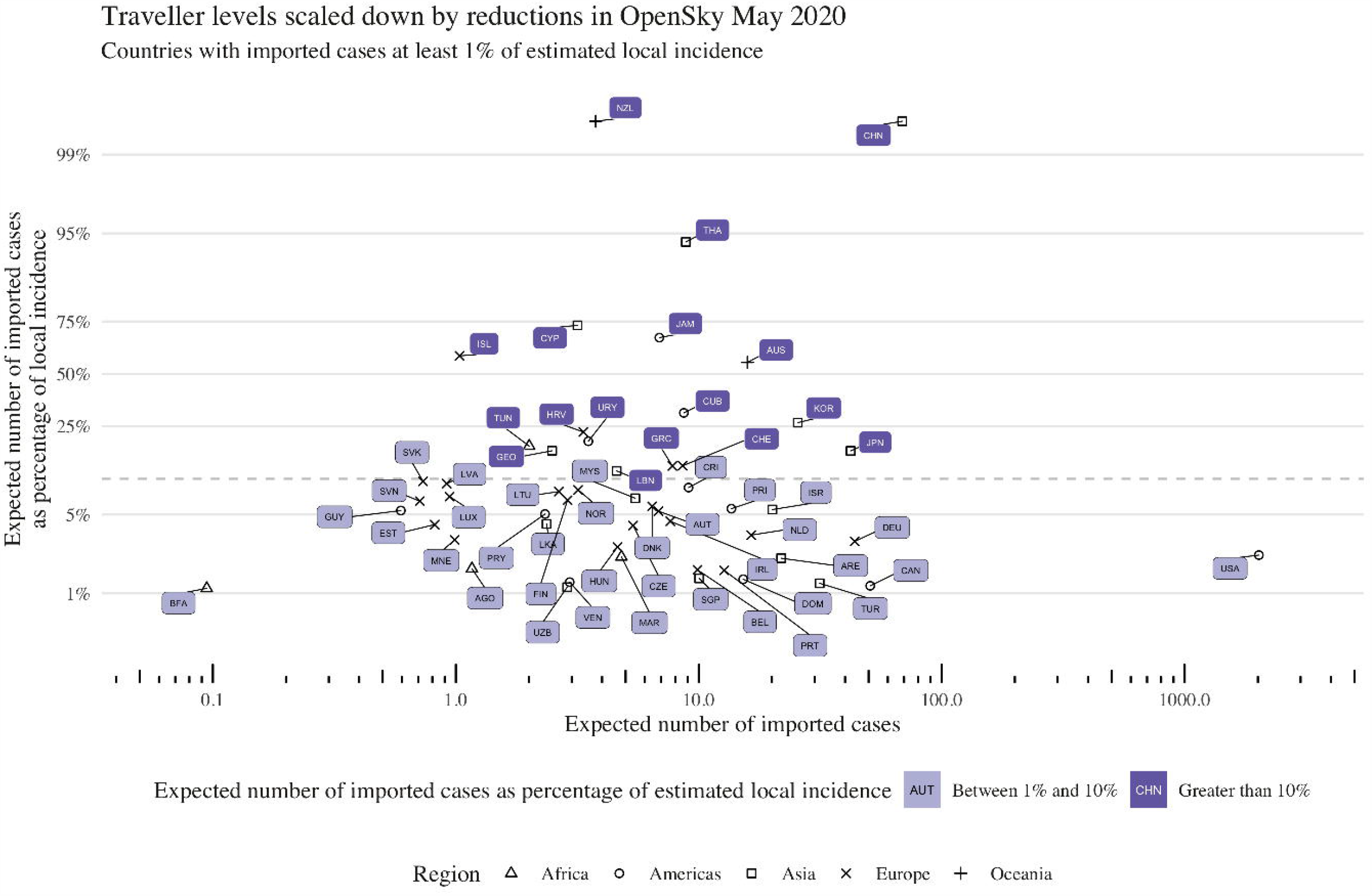
Scatter plot showing the percentage of local daily incidence that daily imported cases represent, where the expected number of imported cases is at least 1% of local incidence. The dashed line represents 10% of local incidence from imported cases. NB: For New Zealand (NZL) and China (CHN), imported cases represent at least 100% of local incidence.

## Discussion

Almost all countries have reported COVID-19 cases, but they differ in the stage of the pandemic they are in (as of July 2020). Many countries in East Asia, Australasia and Europe are well past peak incidence, with some having reduced incidence to very low levels ^10^. Conversely in other countries, incidence remains high and may be increasing. Hence recommendations about international travel restrictions cannot be applied uniformly, but instead need to take into account country circumstances in terms of within-country transmission and connectedness to countries with high prevalence.

Using estimated COVID-19 prevalence in 142 countries together with international travel data between countries, we categorise countries according to the extent to which imported cases may contribute to local transmission in the absence of travel restrictions. In our worst-case scenario (from the perspective of case importation), we assume that travel patterns in May 2019 would hold in May 2020 in the absence of travel restrictions. Even in this scenario, we find that in most countries, imported cases would account for less than 10% of local incidence in the absence of travel restrictions. In other scenarios, we assume that international travel would have decreased in May 2020 (compared to May 2019) even without travel restrictions. In one of these scenarios, we find that in most countries, imported cases would account for less than 10% of local incidence in the absence of travel restrictions.

Hence, our results suggest that in May 2020, travel restrictions may have done little in most countries to change the course of local epidemics, and may not be justified given the high economic and social costs required to prevent the arrival of travellers representing less than 10% (or in many cases, less than 1%) of new cases. In most of the countries where this proportion is greater than 1%, it can be brought below 1% by selective restrictions imposed only on travellers from the highest prevalence countries. However, there are a few countries that would have to prevent entry by almost all international travellers to reach this threshold. These are generally countries where control of local epidemics have been achieved. For instance, both New Zealand and China have low enough incidence that the expected number of imported cases (4 and 69, respectively) is greater than local incidence (2 and 39), so imported cases pose a real risk of triggering a second local epidemic wave (Table S1).

Some countries with moderately large local epidemics in May 2020 (such as some countries in the Americas) still have a moderate level of risk associated with imported cases under the worst-case traveller volume scenario (scenario A), because of their strong connectedness to other high prevalence countries. Imported cases in these countries may be insufficient to drive local epidemics on its own, but may become important in driving epidemics that have already started if countries succeed in reducing local reproduction numbers close to 1, the level at which each new generation of infected cases is smaller than the last.

Our estimates involve simplifying assumptions. We assume that international arrivals in a country have the same probability of being infected as any other person selected at random from the source country. In practice, the risk of infected arrivals is likely to be lower because symptomatic cases are less likely to travel as they may be recuperating at home or in hospital. For those who do attempt to travel, they may be detected during exit screening in the source country or entry screening in the destination country. For those in their incubation period at the time of travel, they may develop symptoms and be detected or sef-declare illness upon arrival. We also did not consider the effect of outbound travellers on local transmission. Travel restrictions would also prevent infected travellers from leaving their source country, which would reduce the number of cases locally, and hence partially mitigate the impact of infected inbound travellers. All these limitations result in overestimating the number of COVID-19 importations that would occur without travel restrictions. However, one limitation in the opposite direction is that we assume that all international travel occurs through flights, so our analysis may not accurately capture the risk of importation between countries that normally have a high volume of land traffic (such as rail and road travel between countries in continental Europe).

Our prevalence and incidence estimates are approximate and may overestimate incidence in countries with younger overall population structures and underestimate it in countries with older populations. ^8^ of these estimates. Furthermore, countries with very low case numbers are excluded from our analysis, as it is not possible to accurately estimate incidence and prevalence estimates for such countries.

Despite these limitations, the categorisation of countries is broadly stable over sensitivity analyses around both country prevalence and incidence estimates, and international travel patterns. They indicate that strict untargeted travel restrictions are probably unjustified in most countries, other than those that have both good international travel connections and very low local COVID-19 incidence. Countries needing to make detailed decisions about travel restrictions or quarantine white lists can use the methods presented here combined with the most current and accurate local data available.

## Data Availability

The data used in this analysis is publicly available case and death time series data, available from the European Centre for Disease Control. The code used for the analysis in this paper is available online (https://github.com/thimotei/covid_travel_restrictions), as is the code used to produce under-reporting estimates, which form the basis for the country-specific prevalence and incidence estimates (https://github.com/thimotei/covid_underreporting).

https://www.ecdc.europa.eu/en/publications-data/download-todays-data-geographic-distribution-covid-19-cases-worldwide

https://github.com/thimotei/covid_underreporting

https://cmmid.github.io/topics/covid19/Under-Reporting.html

https://github.com/thimotei/covid_travel_restrictions

## Contributions

MJ and TWR conceived the study. TWR, JTW and MJ compiled and analysed data. TWR, AJK and WJE estimated country COVID-19 prevalence and incidence. TWR, SC and MJ designed the model and conducted the analyses. All authors contributed to manuscript writing and approved the final version.

## Code and data availability

The data used in this analysis is publicly available case and death time series data, available from the European Centre for Disease Control ^10^. The code used for the analysis in this paper is available online (https://github.com/thimotei/covid_travel_restrictions), as is the code used to produce under-reporting estimates, which form the basis for the country-specific prevalence and incidence estimates (https://github.com/thimotei/covid_underreporting).

## Declaration of interests

We declare no competing interests.

## Acknowledgments

The following funding sources are acknowledged as providing funding for the named authors. MJ was partly funded by the Bill & Melinda Gates Foundation (INV-003174); MJ was partly funded by the National Institute for Health Research (NIHR) using UK aid from the UK Government to support global health research. The views expressed in this publication are those of the author(s) and not necessarily those of the NIHR or the UK Department of Health and Social Care). MJ has received funding from the European Union’s Horizon 2020 research and innovation programme - project EpiPose (101003688: WJE, SC). This project was funded by the Wellcome Trust (206250/Z/17/Z: AJK, 208812/Z/17/Z: SC).

The following funding sources are acknowledged as providing funding for the working group authors. Alan Turing Institute (AE). BBSRC LIDP (BB/M009513/1: DS). This research was partly funded by the Bill & Melinda Gates Foundation (INV-001754: MQ; INV-003174: KP, YL; NTD Modelling Consortium OPP1184344: CABP, GM; OPP1180644: SRP; OPP1183986: ESN; OPP1191821: KO’R, MA). DFID/Wellcome Trust (Epidemic Preparedness Coronavirus research programme 221303/Z/20/Z: CABP, KvZ). DTRA (HDTRA1-18-1-0051: JWR). Elrha R2HC/UK DFID/Wellcome Trust/This research was partly funded by the National Institute for Health Research (NIHR) using UK aid from the UK Government to support global health research. The views expressed in this publication are those of the author(s) and not necessarily those of the NIHR or the UK Department of Health and Social Care (KvZ). ERC Starting Grant (#757688: CJVA, KEA; #757699: JCE, RMGJH; 757699: MQ). This project has received funding from the European Union’s Horizon 2020 research and innovation programme - project EpiPose (101003688: KP, PK, YL). This research was partly funded by the Global Challenges Research Fund (GCRF) project ‘RECAP’ managed through RCUK and ESRC (ES/P010873/1: AG, CIJ, TJ). HDR UK (MR/S003975/1: RME). Nakajima Foundation (AE). NIHR (16/137/109: CD, FYS, YL; 16/137/109 & 16/136/46: BJQ; Health Protection Research Unit for Immunisation NIHR200929: NGD; Health Protection Research Unit for Modelling Methodology HPRU-2012-10096: TJ; PR-OD-1017-20002: AR). Royal Society (Dorothy Hodgkin Fellowship: RL; RP\EA\180004: PK). UK DHSC/UK Aid/NIHR (ITCRZ 03010: HPG). UK MRC (LID DTP MR/N013638/1: GRGL, QJL; MC_PC 19065: RME; MR/P014658/1: GMK). Authors of this research receive funding from UK Public Health Rapid Support Team funded by the United Kingdom Department of Health and Social Care (TJ). Wellcome Trust (206250/Z/17/Z: TWR; 206471/Z/17/Z: OJB; 208812/Z/17/Z: SFlasche; 210758/Z/18/Z: JDM, JH, NIB, SA, SFunk, SRM). No funding (AKD, AMF, DCT, SH).

## List of tables

**Table 1.** Summary of the model parameters. We sum the time to infectiousness and infectious period to arrive at prevalence estimates.

| Model parameter | Description | Value | Source |
| --- | --- | --- | --- |
| Incubation period<br>(days) | Time from exposure to onset of symptoms. | $\Gamma(\mu = 5.5, \sigma^2 = 6.5)$<br>Median: 5.1 days<br>97.5%: 11.5 days | Lauer et al.<sup>13</sup> |
| Time to infectiousness<br>(symptomatic cases) | Time after exposure (and before onset) from which pre-symptomatic transmission can occur. | Median: 3.4 days<br>IQR: (2.3, 4.9) days<br>95%: (0.9, 8.6) days | Derived from He et al.<sup>12</sup> |
| Infectious period<br>(symptomatic cases, days) | Time after incubation period during which case is able to infect others | Median: 7.1 days<br>IQR: (5.7, 8.5) days<br>95%: (2.5, 11.6) days | Derived from Wölfel et al.<sup>14</sup> |

## Notes

### Competing Interest Statement

The authors have declared no competing interest.

### Author Declarations

No exception required

